# The Coronavirus Calendar (CoronaCal): a Simplified SARS-CoV-2 Test System for Sampling and Retrospective Analysis

**DOI:** 10.1101/2022.03.03.22271769

**Authors:** David S. Thaler, Manija A. Kazmi, Karina C. Åberg, Jordan M. Mattheisen, Thomas Huber, Thomas P. Sakmar

## Abstract

**Background:** The testing of saliva samples for severe acute respiratory syndrome coronavirus 2 (SARS-CoV-2) RNA has become a useful and common method to diagnose coronavirus disease 2019 (Covid-19). However, there are limited examples of serial testing with correlated clinical metadata, especially in the outpatient setting.

**Method:** We developed a method to collect serial saliva samples on ordinary white printer paper, which can be subsequently analyzed for the presence of SARS-CoV-2 RNA using established polymerase chain reaction (PCR) procedures. The collection systems consisted of a biological diary (CoronaCal) where subjects dab their saliva onto ovals printed onto paper. The dried samples are covered with a sticker that includes a symptom checklist to create a biological diary. Each sheet of letter paper can accommodate up to 14 serial samples.

**Results:** In a pilot study, ten subjects used CoronaCals for durations of nine to 44 days. SARS-CoV-2 RNA was extracted and detected in CoronaCals from nine of nine people with either Covid-19 symptoms or exposure to someone with Covid-19, and in zero of one asymptomatic person. The CoronaCals were stored for up to 70 days at room temperature during collection and then frozen for up to four months before analysis, suggesting that SARS-CoV-2 RNA is stable once dried onto paper. Interestingly, the temporal pattern of symptoms was not well correlated with SARS-CoV-2 RNA in serial daily collections for up to 44 days. In addition, SARS-CoV-2 positivity was discontinuous over time in most cases but persisted for up to 24 days.

**Conclusions:** We conclude that sampling of saliva on simple paper CoronaCals may provide a useful method to study the natural history and epidemiology of Covid-19. The CoronaCal collection and testing method we developed is also easy to implement, inexpensive, non-invasive, and scalable. More broadly, the approach can be used to archive biological samples for retrospective analysis to deepen epidemiological understanding during viral disease outbreaks and to provide information about the natural history of emerging infections.

## INTRODUCTION

Insights enabled by after-action and historical analysis of infectious diseases depend importantly on preserved samples both biological and bio-cultural [1-3]. If properly documented samples were widely saved, then as resource allocations allow, key parameters of pandemic outbreaks might become better understood. For example, in cases of emerging viral diseases, knowledge about incubation periods, the relationship between transmissibility and symptoms, and variability in the length of time that individuals stay infected could be used to plan effective mitigation strategies [4].

Nucleic acid testing for SARS-CoV-2 is currently limited on three fronts: 1) sample collection, 2) transportation and processing of samples, 3) running assays, recording, and reporting results in a timely manner [5]. Rapid real-time testing is presently prioritized based on urgent and immediate needs. Primary biological samples may be preserved in liquids, indefinitely, at room temperature [6, 7]. However, storage of samples kept in this way requires space and organizational resources that often cannot be prioritized either in acute circumstances or as a long-term commitment of resources. Isolated nucleic acids are stable at room temperature, but purification requires significant resources. Previous work has shown that nucleic acids, DNA and RNA, are recoverable from biological liquids dried on specialized paper filters and stored in freezers [8]. Saliva is easy to sample and widely used for clinical SARS-CoV-2 real-time quantitative (RT-q) PCR assays [9-11].

Here, we tested the hypothesis that saliva dried onto ordinary laser printer paper and stored at either room temperature and/or in a household freezer preserves RNA, allowing for later recovery and assay. We designed and implemented a biological diary called CoronaCal to allow for simple daily saliva self-collection in real-life situations in the community. SARS-CoV-2 RNA was transiently detected in volunteers who contemporaneously recorded symptoms over many days, which coincided with virus infection as shown by conventional assays. Human RNase P RNA was used as a positive control and was shown to be stable over several months. The ability to assay SARS-CoV-2 RNA on archive calendars has important potential to enrich epidemiology and thereby contribute to rational implementation of control measures.

## MATERIALS AND METHODS

### Apparatus

The Agilent AriaMx qPCR instrument was used for RT-qPCR measurements.

### Materials and reagents

RNA extraction from the CoronaCals were carried out using Trizol reagent from Life Technologies (Cat#15596026) and the Direct-Zol RNA Microprep kit from Zymo Research (Cat#3R2060). Elution was in molecular biology grade water containing RNase Inhibitor New England Biolabs (cat#0314S). Real time RT-qPCR was carried out using the GoTaq Probe 1 Step RT-qPCR system from Promega (cat#A6120). All primers, probes and control plasmids were obtained from IDT Integrated DNA Technologies; N1 and N2 primer probes from the 2019 nCoV CDC EUA Kit (cat#10006606), RP primers RNase P Forward (cat#10006827), RNase P Reverse (cat#10006828) and Rnase P ATTO 647 probe (cat#10007061). Positive control plasmids, IDT 2019 nCoV_N_Positive Control 200,000 copies/uL and IDT 2019 Hs_RPP30_Positive Control 200,000 copies/uL.

### Sample collection

CoronaCal was designed to be used by anonymous, unidentified volunteer participants without counseling. The paper sampling diaries were printed onto Hammermill ForeR Multi-Purpose 20LB white letter-sized printer paper. The stickers used as symptom log lists were printed on Avery 1” x 2-5/8” Rectangle 6460 labels using a Hewlett-Packard Color LaserJet 4700DN printer (see Supplemental Figure 1). The materials were packaged along with an instruction sheet into a tear-resistant envelope, which was also used to return the completed CoronaCals. Volunteer participants from the community were asked to prepare a CoronaCal if they had either tested positive on a PCR-Covid-19 test or had been exposed to someone who had recently tested positive.

The participants remained anonymous and unidentified, and data obtained from the diaries was not correlated with any information external to what was archived on the CoronaCals themselves. The protocol for the collection of the biological diaries described in this study was reviewed by the Institutional Review Board (IRB) at Rockefeller University and was deemed not to be human-subjects research.

Participants were instructed to wash their hands, and then with a clean finger apply a dab of saliva to coat completely the oval corresponding to the collection day on the CoronaCal. While the saliva air dried, the participants filled out the survey on the Avery label with symptoms for that day and then used it to seal the oval containing that day’s dried saliva sample. CoronaCals were kept at room temperature for duration of collection period then returned to the laboratory where they were stored for up to four months at -20°C until assayed.

### Sample processing

The CoronaCals were processed as follows: Eight 2-mm-wide x ∼17-mm long paper strips were excised in a multiplexed fashion from the oval collection zone for each day on the diary using a concatenated razor blade device. After removing the adhesive, each strip was transferred into RNase/DNase free 1.5-mL tubes. We estimate that the equivalent of about 3-5 μL of saliva was present on each strip of paper cut from the sample application oval.

### RNA Extraction and purification

Each sample was incubated in 300 μL of Trizol for 15 min at room temperature while shaking at 500 rpm. Next, RNA from the sample was purified using the Zymo Direct-zol RNA microprep kit following manufacturer’s protocol and eluted with 15 μL of DNase/RNase-Free molecular biology grade water containing RNase inhibitor (17 units).

### SARS-CoV-2 detection

SARS-CoV-2 RNA was detected using Promega’s GoTaq Probe 1 Step RT-qPCR system. Reactions were set up in duplicate in 96-well plates using 3 μL of template generated above multiplexed with either the N1 or N2 primer probe from IDT 2019 nCoV CDC EUA Kit along with the RP primer set and ATTO probe. Reactions were run on the Agilent AriaMx Real-time PCR System with the following cycling conditions: Reverse transcription 15 min, 45°C; hot start 2 min, 95°C; amplification 45 cycles 3 sec, 95°C, 30 sec 55°C. Each plate had a multiplex control reaction containing 2,000 copies each of IDT 2019 nCoV_N_Positive Control and IDT 2019 Hs_RPP30_Positive Controls. Data were recorded for up to 45 PCR cycles to give a maximum cycle threshold (Ct) of 45 on data plots (Figure 1).

**Figure 1.**
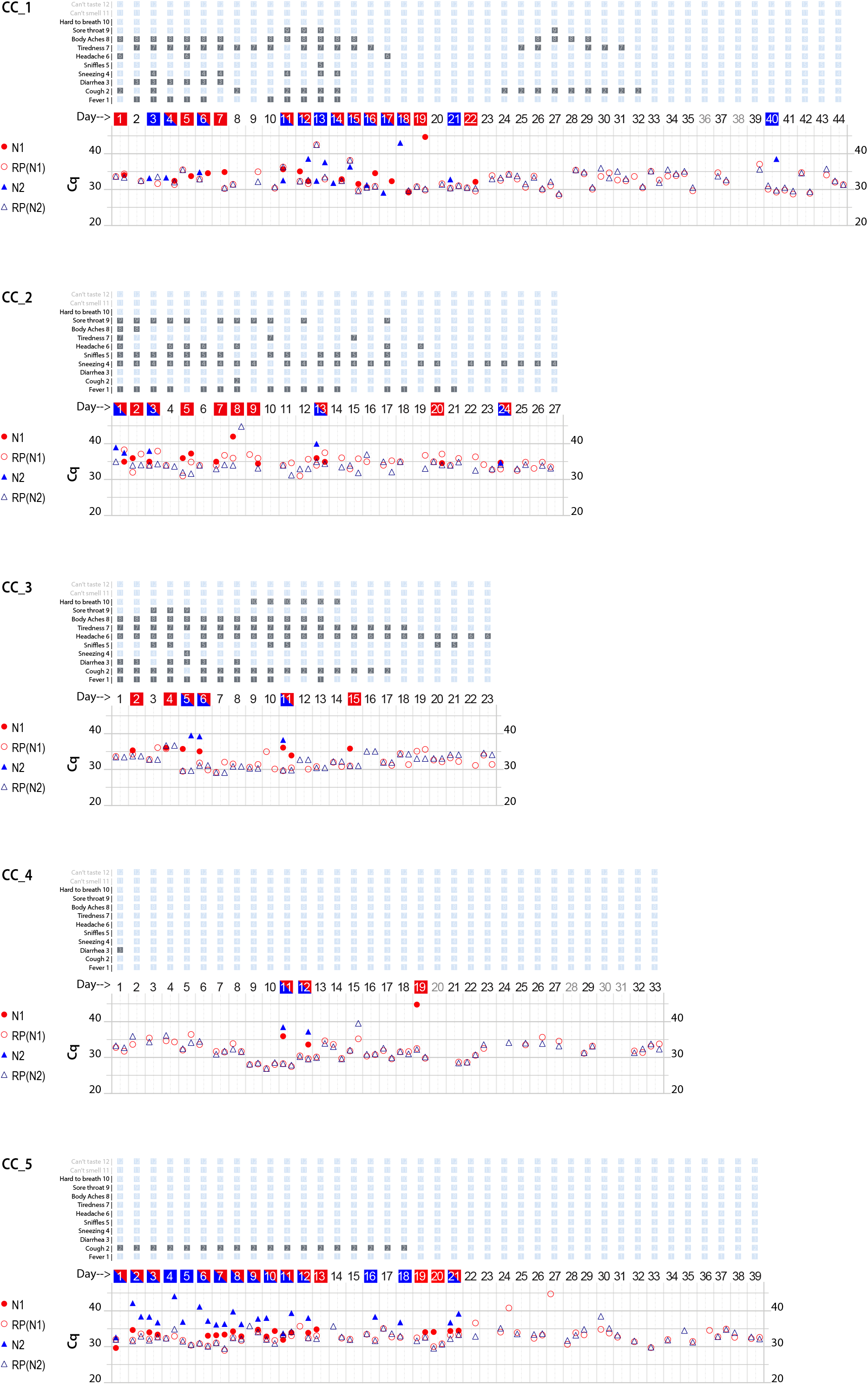
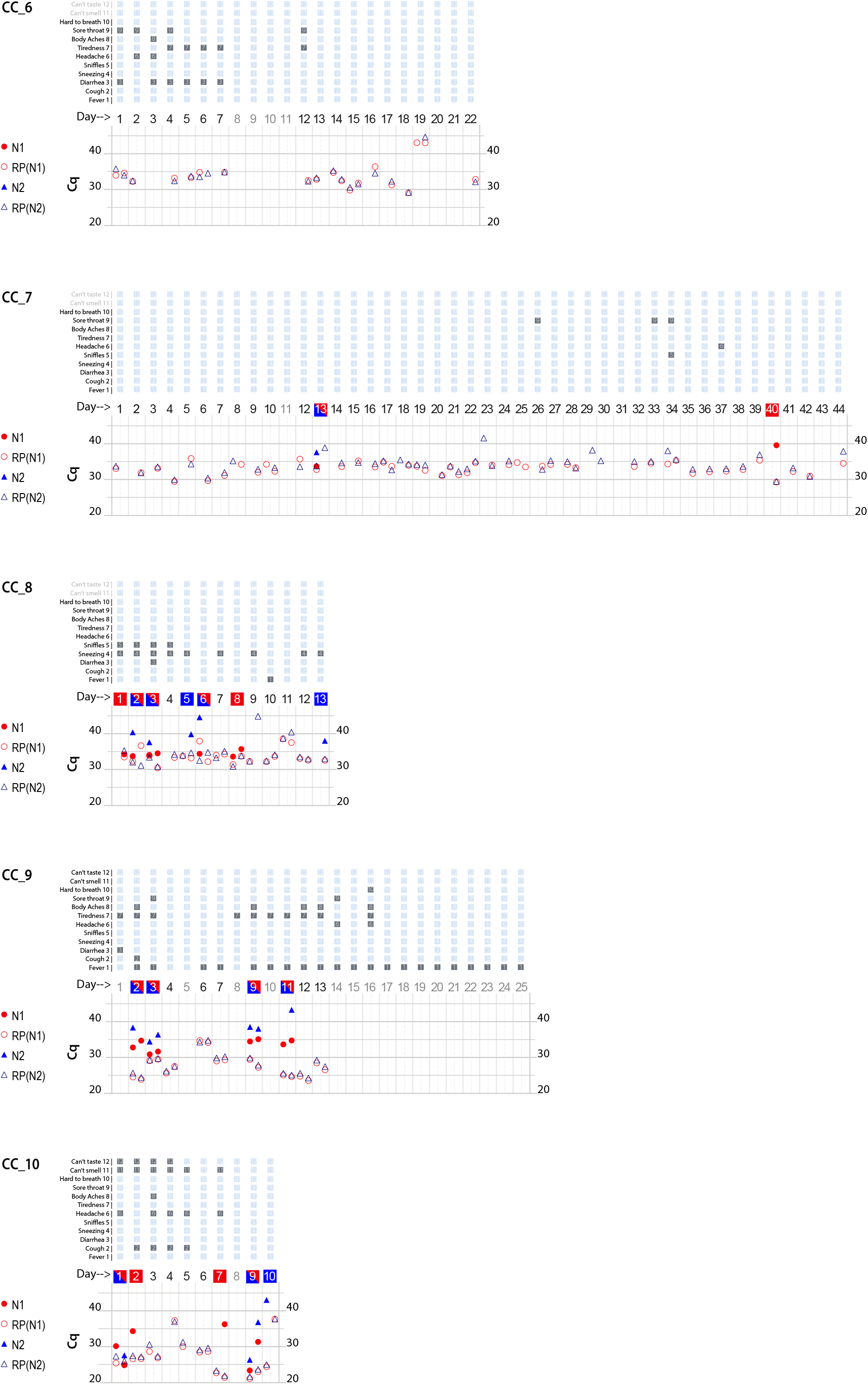
Graphical presentation of data obtained from ten CoronaCal diaries. The ten CoronaCals presented are labeled CC_1 through CC_10. For example, CC_1 shows data from a CoronaCal diary in which one individual collected saliva samples and self-recorded symptoms for 44 consecutive days during the course of a Covid-19 illness. Each chart is essentially a timeline that can be read from left to right. Each column demarked by vertical grey lines represents one day. At the top of the figure, filled boxes represent self-reported symptoms: 1, fever; 2, cough; 3, diarrhea; 4, sneezing; 5, sniffles; 6, headaches; 7, tiredness; 8, body aches; 9, sore throat; 10, hard to breath; 11, can’t smell; 12, can’t taste. The bottom section of the chart shows the results of RT-qPCR analysis of RNA samples extracted from the paper CoronaCal. Duplicate samples were analyzed for each day using PCR primer/probe combinations designed to detect SARS-CoV-2 N1 (red solid circles) and N2 (blue solid triangles). For each sample, and for each N1/N2 probe, saliva RNAase P RNA was measured as a control (open circles and triangles). The day numbers are boxed in red when the PCR threshold for N1 positive was reached, blue if the Cq threshold for N2 positive was reached, or both red and blue when both N1 and N2 were deemed to be positive. An absence of symbols indicates that no amplicon was detected after 45 PCR cycles, and days in light grey indicate days where no sample was collected.

### Testing limit of detection

Additionally, we tested the limit of detection and linearity of the SARS-CoV-2 virus RNA assay using the control plasmids IDT 2019 nCoV_N_Positive Control and IDT 2019 Hs_RPP30_Positive Control. Plasmids were serially diluted from 100,000 copies per reaction down to 0.1 copies per reaction in water and RT-qPCR reactions were set up in triplicate and performed as described above to determine the Ct values for each dilution. The results are shown in Supplementary Figure 2.

#### Instruments and Reagents

*Hammermill ForeR Multi-Purpose 20LB white printer paper*

*Avery 1” x 2-5/8” Rectangle 6460 labels, removable matte white paper*

*Hewlett-Packard Color LaserJet 4700DN printer*

*GoTaq Probe 1 Step RT-qPCR system Promega A6120*

*IDT 2019 nCoV CDC EUA Kit #10006606 (IDT Integrated DNA Technologies)*

*IDT 2019 nCoV_N_Positive Control 200,000 copies/*μL

*IDT 2019 Hs_RPP30_Positive Control 200,000 copies/*μL

*Trizol reagent Life Technologies #15596026 100ml*

*Zymo Research Direct-Zol RNA Microprep 3R2060*

*New England Biolabs RNase Inhibitor 0314S (40units/μL)*

#### Consumables

*AriaMx 96-well plates VWR Agilent #401490*

*Adhesive seals Agilent #401492*

*Applied Biosystems optical cover compression pad, Part#4312639*

*Qiagen VacConnectors (500) Cat#19407*

#### RT-qPCR Primers

*RP primer and ATTO probe (IDT Integrated DNA Technologies)*

*RNase P Forward (IDT 10006827), RNase P Reverse (IDT 10006828) Rnase P ATTO 647 (IDT 10007061)*

*20μL RP forward, 20μL RP reverse, 10μL RP probe, 950μL TE*

*-use 2μL per reaction*

*N1 and N2 primer probe from IDT 2019 nCoV CDC EUA Kit #10006606*

*-use 1*.*5μL each per reaction*

#### Controls

*IDT 2019 nCoV_N_Positive Control 200,000 copies/μL*

*IDT 2019 Hs_RPP30_Positive Control 200,000 copies/μL*

*-Make a stock containing 500 copies/μL of each control. Use 3μL per reaction*.

#### RT-qPCR

*GoTaq Probe 1 Step RT-qPCR system - Prepare the 1X master mix containing carboxy-X-rhodamine (CXR) reference dye for N1+RP, and N2+RP primer/probe and GoScript and dispense 17μL per well of a 96-well plate. Use 3μL template per reaction. Seal plate and give a quick vortex and spin, place an optical cover compression pad over plate and place into AriaMx*.

|  |  |  |  |
| --- | --- | --- | --- |
| *RT* | *45°C* | *15 min* | *1 cycle* |
| *Hot start* | *95°C* | *2 min* | *1 cycle* |
| *Amplification* | *95°C* | *3 sec/55°C 30 sec* | *45 cycles* |

## RESULTS AND DISCUSSION

The CoronaCal diary presented to participants is shown in Supplemental Figure 1. Volunteers self-sampled and kept their CoronaCals at home at room temperature during their entire sampling period. Once the sample period was completed, CoronaCals were stored at -20°C prior to sample extraction for up to four months.

RNase P and SARS-CoV-2 RNA signals were reproducibly detected from multiple samplings after different durations of storage. The longest CoronaCal included samples from 44 consecutive days. No signal diminution was seen when testing samples stored at different durations at - 20°C, or after multiple freeze-thaw cycles. These results suggest that the RNA in the dried saliva samples is stable at room temperature for weeks, and indefinitely at -20°C. The sensitivity of the RT-qPCR is given in Supplementary Figure 2.

This pilot study reports results from CoronaCals processed and analyzed from ten individuals. As shown in Figure 1, each CoronaCal is the record of a different individual anonymous volunteer. Nucleic acids were extracted from CoronaCals and assayed by reverse transcriptase RT-qPCR as described in Materials and Methods. All assays were carried out in duplicate. Greater than ∼90% of the time duplicates were in close concordance. In the few exceptions where only one of two of the replicates gave a positive signal, it was still recorded and included in the data plots.

CoronaCals 1 (CC_1) and 7 (CC_7) were sampled for 44 continuous days (with the exceptions of a few blank days on CC_1). The control signal of human RNase P was consistent across the entire time series in these samples. This serves as a check for the assay procedure, including the extraction and RT-qPCR steps. Importantly, these results show the RNA moiety of human RNase P was stable under these conditions because the quantification PCR cycle threshold (Cq) does not increase across the sample. If RNA target degradation were occurring on the stored CoronaCal, one might expect that samples earlier in the series would have less signal than those later in the series. Samples from day one, were *in situ* and at room temperature on the CoronaCal for between 5 and 6 weeks longer than the last samples collected. Detection of RNase P was reliable and continuous over the entire period showing the stability at room temperature of the RNA signal under these conditions.

CoronaCals were printed on ordinary paper with designated spaces to place a fingertip’s worth of saliva (∼ 30-50 μL) each day. The saliva spot was allowed to air dry, then covered with an adhesive label. The covering label serves two functions: 1) it isolates samples preventing cross contamination b) it decreases the already very small chance of fomite transmission [18] or aerosolization from dried saliva. The covering sticker was also a label with space for recording symptoms and could conceivably be used for a barcode that could be coordinated with cellphone apps for location, proximity to others, and analysis pipelines. The paper preservation format could be implemented by individuals or employed in a coordinated way for groups traveling, working, or living together.

Each CoronaCal relates the complete story of the course of one person’s Covid-19 illness. It is instructive to describe the results of several of the CoronaCals in detail. For example, CC_1 reports over a 44-day period. On Day 1, the subject reported cough, headache, and body aches and was also positive for SARS-CoV-2 RNA (N1 only) in their saliva. On Day 3 through Day 7 of illness, the positive saliva RNA persisted (only one of the RNase P controls was positive on Day 2), and the symptoms changed. Fever appeared on Day 2 along with other symptoms. By Day 8 of the illness, some symptoms had subsided, and there was no longer SARS-CoV-2 RNA present in the saliva. However, on Day 11, after three consecutive days with fewer symptoms and no detectable SARS-CoV-2 RNA in the saliva, fever recurred, as did the appearance of SARS-CoV-2 RNA in the saliva on Day 11. On Day 18 of the illness, the subject was asymptomatic, but continued to be positive for saliva RNA for four out of the next five days.

SARS-CoV-2 positive RT-qPCR signals, when they appeared, were transient and discontinuous, consistent with the natural course of infection as seen in studies that conducted standard qPCR assays each day [12]. None of the nine CoronaCals with positive results for SARS-CoV-2 RNA detection had continuous daily positive tests, became negative and then remained negative. All the positives were discontinuously positive. The participant who was positive for SARS-CoV-2 RNA for the longest duration was CC_5, who was positive for 13 consecutive days. CC_5 was also positive for 18 of the first 21 days of illness. CC_5 reported only cough, but for 18 consecutive days, and also remained positive for SARS-CoV-2 RNA even after becoming asymptomatic. The duration of positive tests was striking with CC_2, which showed positive tests for ten days over a span of 24 days (see also CC_1 and CC_5). One caveat is that not all testing detected both N1 and N2. In addition, the presence of small amounts of detectable SARS-CoV-2 RNA does not confirm the presence of infectious viral particles.

The relationship between self-reported symptoms and SARS-CoV-2 RNA signal is of interest. Large studies have shown that positive PCR tests are not always associated with symptoms and conversely, that symptoms can occur during the absence of a PCR signal [12]. The CoronaCals presented here demonstrate these possibilities directly during the time courses of infections in individuals.

The CoronaCal method relies totally on self-reporting and self-collection. The study team had no direct contact with the volunteer participants to instruct or influence their use of the diaries or to attempt to increase compliance. Although participants were provided with enough materials to collect up to 84 days of samples, the longest diaries collected were from 44 days. The shortest diary collected was for 10 days. Compliance is an issue for any self-collection and reporting system, and although compliance was not uniform, a tremendous amount of information could be obtained if the strategy were to be scaled-up to larger numbers of participants. In addition, more information could be obtained if participants could be interviewed, or if everyone in a particular household, for example, completed a CoronaCal diary contemporaneously. Such a study would require informed consent, however, and was not our aim for a proof-of-concept pilot study to develop and validate the methodology.

CoronaCals CC_1 through CC_8 were collected from participants in New York City during March through June 2020 when the original Wuhan SARS-CoV-2 was prevalent. CC_9 and CC_10 were collected later when the so-called Alpha variant was dominant. All ten CoronaCals were collected before COVID-19 vaccines were available, so none of the participants were vaccinated. The underlying health status or Covid-19 risk factors are not known for any of the anonymous and unidentified participants.

A key and surprising finding of this work is that specific RNA signals for SARS-CoV-2 as well as human RNase P were stable for extended periods in small samples of saliva dried onto ordinary printer paper. This finding was used to design a convenient user-friendly format for individuals to collect and preserve personal daily samples of oral biological fluids. Saliva samples, dried down onto a calendar printed on ordinary printer paper became a biological diary that supported molecular analysis weeks or months after the samples were deposited. This work is a proof of principle. It was not a forgone conclusion that nucleic acids, in particular RNA, would be stable enough to support detection after extended periods on commonplace substrates at room temperature and standard -20°C freezer storage.

The stability of RNA signal is evident in the consistency and steadiness of RNase P control detection. For example, from CC_1 the control RNA Cq values are not significantly different on Day 1 than on Day 44 (the average Cq = 32.7 +/- 2.4 S.D. for the RNase P control signal). Even though Cq values were consistent for a given CoronaCal, they did seem to vary when averages were compared among the ten CoronaCals tested, suggesting that individuals dabbed different amounts of saliva, but each individual was consistent in how much they dabbed from one day to the next. The Day 1 sample for CC_1 was kept at room temperature for at least 44 days before it was placed at -20°C for long-term storage. Of course, day-to-day Cq controls are affected by the amount of saliva applied, which is not measured or so controlled, and is therefore expected to vary.

The goal of this study was to find the right balance between reproducibility and ease-of-use. The stability of RNA signals found in this study is conceivably due to special features of the RNAs involved. SARS-CoV-2 RNA packaged inside virions may be protected from nucleases and/or otherwise stabilized [13]. Having said that, it is not known from this work, or from other studies of which we are aware, how much of the SARS-CoV-2 signal detected in saliva is present in packaged virions, and how much is inside epithelial cells that have been shed into saliva. RNase P RNA is normally part of a large multi-subunit complex, and this complex might shield the RNA signal [14, 15]. At this time, the stability of other RNA moieties in this format, such as mRNA, is unknown. Our hypothesis is that rapid drying saliva samples onto a porous paper surface is the key factor responsible for signal stability.

Previous studies have shown that SARS-CoV-2 RNA is detectable for extended periods on surfaces [16]. The present work exploits this observation to develop a format for non-invasive, resource-sparing sample archiving, extends the sampling period, and uses ordinary printer paper. In addition, the distinction between detectable RNA and infective virus is important to keep in mind, since virions can be rendered non-infectious before RNA becomes non-detectable [17].

The function of retrospective assay from dried samples stored for long periods should not be confused with the requirements of fresh clinical samples whose purpose is rapid diagnosis. Physical anthropologists, forensic practitioners, and historical epidemiologists often obtain valuable data from materials that do not meet the standards of clinical diagnosis. Most nucleic acid recovery from non-optimal and ancient materials has been DNA [19]. However, the recovery of RNA sequences from the 1918-1919 influenza via paraffin sections and victims exhumed from permafrost graves [20] and HIV from lymph node embeds from 1960s [21] shows that RNA under at least some circumstances is more stable than often assumed.

Archived samples have previously proven important in tracing the origin and spread of infectious diseases. However, in the cases of which we are aware, materials were originally collected for other purposes and never systematic time series. Molecular analysis used to study the origin of HIV did not exist at the time samples were taken [21]. Archived biological diaries will allow systematic study of infectious history and personalized medicine whose potential uses extend beyond the targets examined in this study.

The present work focuses on SARS-CoV-2, but clearly there are further applications. Host and microbiome nucleic acids as well as other biomarkers are all of possible interest. Some assays would be specific for already-known targets. Discovery approaches such as untargeted sequencing and other “omics” have the potential to identify novel infectious agents and biomarkers beyond those that have been characterized at the time of sample collection [22]. Future developments of analytical methods may reasonably be predicted to increase the value of archived biological samples, and samples are expected to contain more potential information than can currently be interpreted [23]. In addition to records of individuals, related approaches would be valuable if applied to wastewater [24, 25] and eco-environmental contexts. Archived biological diaries from multiple levels would add rich information to retrospective reviews aimed at improving pandemic response [26], public health more generally[27] and enhancing understanding of our relentlessly evolving world[19, 28, 29].

CoronaCal style diaries could be widely used and provide valuable information on individuals, and communities to provide valuable cumulative data to better understand pathways of contagion and effectiveness of containment approaches. Biological diaries such as CoronaCal may also be valuable in contexts of individualized medicine and to better understand and thus optimize public health measures. Our hope is that others will adopt the use of the CoronaCal strategy to study Covid-19 in their own communities and to archive samples for future study.

## Data Availability

All data produced in the present study are available upon reasonable request to the authors.

https://www.coronacal.com

## ACKNOWLEDGMENTS

We are grateful for financial support from the Richard Lounsbery Foundation and The Rockefeller University. We wish to thank Jesse Ausubel, Richard P. Lifton, Tom Tuschl, Ann H. Campbell, Ashley Foo, and Chris Keough. This work was inspired by our involvement with the Leonardo Da Vinci DNA Project, and we thank all project members for valuable discussions.

