## Supplemental material for "The Coronavirus Calendar (CoronaCal): a Simplified SARS-CoV-2 Test System for Sampling and Retrospective Analysis"

Running title:
SARS-CoV-2 sample preservation on paper diaries

**CoronaCal™** *Use a clean fingertip to fill the oval with your saliva. When it has dried, align today's sticker within the dotted line. It will cover this message and the DRIED saliva. Check the boxes relevant to your symptoms. Thank you for your contribution!*

### WEEK #1

*Use a clean fingertip to fill the oval with your saliva. When it has dried, align today's sticker within the dotted line: It will cover this message and the DRIED saliva. Check the boxes relevant to your symptoms. Thank you!*

Day 1

*Use a clean fingertip to fill the oval with your saliva. When it has dried, align today's sticker within the dotted line: It will cover this message and the DRIED saliva. Check the boxes relevant to your symptoms. Thank you!*

Day 2

*Use a clean fingertip to fill the oval with your saliva. When it has dried, align today's sticker within the dotted line: It will cover this message and the DRIED saliva. Check the boxes relevant to your symptoms. Thank you!*

Day 3

*Use a clean fingertip to fill the oval with your saliva. When it has dried, align today's sticker within the dotted line: It will cover this message and the DRIED saliva. Check the boxes relevant to your symptoms. Thank you!*

Day 4

*Use a clean fingertip to fill the oval with your saliva. When it has dried, align today's sticker within the dotted line: It will cover this message and the DRIED saliva. Check the boxes relevant to your symptoms. Thank you!*

Day 5

*Use a clean fingertip to fill the oval with your saliva. When it has dried, align today's sticker within the dotted line: It will cover this message and the DRIED saliva. Check the boxes relevant to your symptoms. Thank you!*

Day 6

*Use a clean fingertip to fill the oval with your saliva. When it has dried, align today's sticker within the dotted line: It will cover this message and the DRIED saliva. Check the boxes relevant to your symptoms. Thank you!*

Day 7

### WEEK #2

*Use a clean fingertip to fill the oval with your saliva. When it has dried, align today's sticker within the dotted line: It will cover this message and the DRIED saliva. Check the boxes relevant to your symptoms. Thank you!*

Day 8

*Use a clean fingertip to fill the oval with your saliva. When it has dried, align today's sticker within the dotted line: It will cover this message and the DRIED saliva. Check the boxes relevant to your symptoms. Thank you!*

Day 9

*Use a clean fingertip to fill the oval with your saliva. When it has dried, align today's sticker within the dotted line: It will cover this message and the DRIED saliva. Check the boxes relevant to your symptoms. Thank you!*

Day 10

*Use a clean fingertip to fill the oval with your saliva. When it has dried, align today's sticker within the dotted line: It will cover this message and the DRIED saliva. Check the boxes relevant to your symptoms. Thank you!*

Day 11

*Use a clean fingertip to fill the oval with your saliva. When it has dried, align today's sticker within the dotted line: It will cover this message and the DRIED saliva. Check the boxes relevant to your symptoms. Thank you!*

Day 12

*Use a clean fingertip to fill the oval with your saliva. When it has dried, align today's sticker within the dotted line: It will cover this message and the DRIED saliva. Check the boxes relevant to your symptoms. Thank you!*

Day 13

*Use a clean fingertip to fill the oval with your saliva. When it has dried, align today's sticker within the dotted line: It will cover this message and the DRIED saliva. Check the boxes relevant to your symptoms. Thank you!*

Day 14

☐ Fever
☐ Cough
☐ Diarrhea
☐ Sneezing
☐ Sniffles
☐ Headache
☐ Tiredness
☐ Body aches
☐ Sore throat
☐ Hard to breathe
CoronaCal

☐ Fever
☐ Cough
☐ Diarrhea
☐ Sneezing
☐ Sniffles
☐ Headache
☐ Tiredness
☐ Body aches
☐ Sore throat
☐ Hard to breathe
CoronaCal

☐ Fever
☐ Cough
☐ Diarrhea
☐ Sneezing
☐ Sniffles
☐ Headache
☐ Tiredness
☐ Body aches
☐ Sore throat
☐ Hard to breathe
CoronaCal

☐ Fever
☐ Cough
☐ Diarrhea
☐ Sneezing
☐ Sniffles
☐ Headache
☐ Tiredness
☐ Body aches
☐ Sore throat
☐ Hard to breathe
CoronaCal

☐ Fever
☐ Cough
☐ Diarrhea
☐ Sneezing
☐ Sniffles
☐ Headache
☐ Tiredness
☐ Body aches
☐ Sore throat
☐ Hard to breathe
CoronaCal

☐ Fever
☐ Cough
☐ Diarrhea
☐ Sneezing
☐ Sniffles
☐ Headache
☐ Tiredness
☐ Body aches
☐ Sore throat
☐ Hard to breathe
CoronaCal

☐ Fever
☐ Cough
☐ Diarrhea
☐ Sneezing
☐ Sniffles
☐ Headache
☐ Tiredness
☐ Body aches
☐ Sore throat
☐ Hard to breathe
CoronaCal

☐ Fever
☐ Cough
☐ Diarrhea
☐ Sneezing
☐ Sniffles
☐ Headache
☐ Tiredness
☐ Body aches
☐ Sore throat
☐ Hard to breathe
CoronaCal

☐ Fever
☐ Cough
☐ Diarrhea
☐ Sneezing
☐ Sniffles
☐ Headache
☐ Tiredness
☐ Body aches
☐ Sore throat
☐ Hard to breathe
CoronaCal

☐ Fever
☐ Cough
☐ Diarrhea
☐ Sneezing
☐ Sniffles
☐ Headache
☐ Tiredness
☐ Body aches
☐ Sore throat
☐ Hard to breathe
CoronaCal

☐ Fever
☐ Cough
☐ Diarrhea
☐ Sneezing
☐ Sniffles
☐ Headache
☐ Tiredness
☐ Body aches
☐ Sore throat
☐ Hard to breathe
CoronaCal

☐ Fever
☐ Cough
☐ Diarrhea
☐ Sneezing
☐ Sniffles
☐ Headache
☐ Tiredness
☐ Body aches
☐ Sore throat
☐ Hard to breathe
CoronaCal

☐ Fever
☐ Cough
☐ Diarrhea
☐ Sneezing
☐ Sniffles
☐ Headache
☐ Tiredness
☐ Body aches
☐ Sore throat
☐ Hard to breathe
CoronaCal

☐ Fever
☐ Cough
☐ Diarrhea
☐ Sneezing
☐ Sniffles
☐ Headache
☐ Tiredness
☐ Body aches
☐ Sore throat
☐ Hard to breathe
CoronaCal

☐ Fever
☐ Cough
☐ Diarrhea
☐ Sneezing
☐ Sniffles
☐ Headache
☐ Tiredness
☐ Body aches
☐ Sore throat
☐ Hard to breathe
CoronaCal

☐ Fever
☐ Cough
☐ Diarrhea
☐ Sneezing
☐ Sniffles
☐ Headache
☐ Tiredness
☐ Body aches
☐ Sore throat
☐ Hard to breathe
CoronaCal

☐ Fever
☐ Cough
☐ Diarrhea
☐ Sneezing
☐ Sniffles
☐ Headache
☐ Tiredness
☐ Body aches
☐ Sore throat
☐ Hard to breathe
CoronaCal

☐ Fever
☐ Cough
☐ Diarrhea
☐ Sneezing
☐ Sniffles
☐ Headache
☐ Tiredness
☐ Body aches
☐ Sore throat
☐ Hard to breathe
CoronaCal

☐ Fever
☐ Cough
☐ Diarrhea
☐ Sneezing
☐ Sniffles
☐ Headache
☐ Tiredness
☐ Body aches
☐ Sore throat
☐ Hard to breathe
CoronaCal

☐ Fever
☐ Cough
☐ Diarrhea
☐ Sneezing
☐ Sniffles
☐ Headache
☐ Tiredness
☐ Body aches
☐ Sore throat
☐ Hard to breathe
CoronaCal

☐ Fever
☐ Cough
☐ Diarrhea
☐ Sneezing
☐ Sniffles
☐ Headache
☐ Tiredness
☐ Body aches
☐ Sore throat
☐ Hard to breathe
CoronaCal

☐ Fever
☐ Cough
☐ Diarrhea
☐ Sneezing
☐ Sniffles
☐ Headache
☐ Tiredness
☐ Body aches
☐ Sore throat
☐ Hard to breathe
CoronaCal

☐ Fever
☐ Cough
☐ Diarrhea
☐ Sneezing
☐ Sniffles
☐ Headache
☐ Tiredness
☐ Body aches
☐ Sore throat
☐ Hard to breathe
CoronaCal

☐ Fever
☐ Cough
☐ Diarrhea
☐ Sneezing
☐ Sniffles
☐ Headache
☐ Tiredness
☐ Body aches
☐ Sore throat
☐ Hard to breathe
CoronaCal

☐ Fever
☐ Cough
☐ Diarrhea
☐ Sneezing
☐ Sniffles
☐ Headache
☐ Tiredness
☐ Body aches
☐ Sore throat
☐ Hard to breathe
CoronaCal

☐ Fever
☐ Cough
☐ Diarrhea
☐ Sneezing
☐ Sniffles
☐ Headache
☐ Tiredness
☐ Body aches
☐ Sore throat
☐ Hard to breathe
CoronaCal

☐ Fever
☐ Cough
☐ Diarrhea
☐ Sneezing
☐ Sniffles
☐ Headache
☐ Tiredness
☐ Body aches
☐ Sore throat
☐ Hard to breathe
CoronaCal

☐ Fever
☐ Cough
☐ Diarrhea
☐ Sneezing
☐ Sniffles
☐ Headache
☐ Tiredness
☐ Body aches
☐ Sore throat
☐ Hard to breathe
CoronaCal

☐ Fever
☐ Cough
☐ Diarrhea
☐ Sneezing
☐ Sniffles
☐ Headache
☐ Tiredness
☐ Body aches
☐ Sore throat
☐ Hard to breathe
CoronaCal

☐ Fever
☐ Cough
☐ Diarrhea
☐ Sneezing
☐ Sniffles
☐ Headache
☐ Tiredness
☐ Body aches
☐ Sore throat
☐ Hard to breathe
CoronaCal

Supplemental Figure 1. **The CoronaCal diary.** *Upper panel.* CoronaCal collection sheets were printed onto ordinary printer paper. Each 8-1/2" x 11" sheet can accommodate two-weeks of daily samples, and each participant was provided with enough sheets for up to 12 weeks of sampling. *Lower panel.* Covering stickers with a symptom check list were printed on Avery sticker stock. A later version of the symptoms list included "can't smell" and "can't taste." The collection sheet and sticker templates are reduced here from their actual size. Downloadable CoronaCal templates are available www.CoronaCal.com as a supplement.

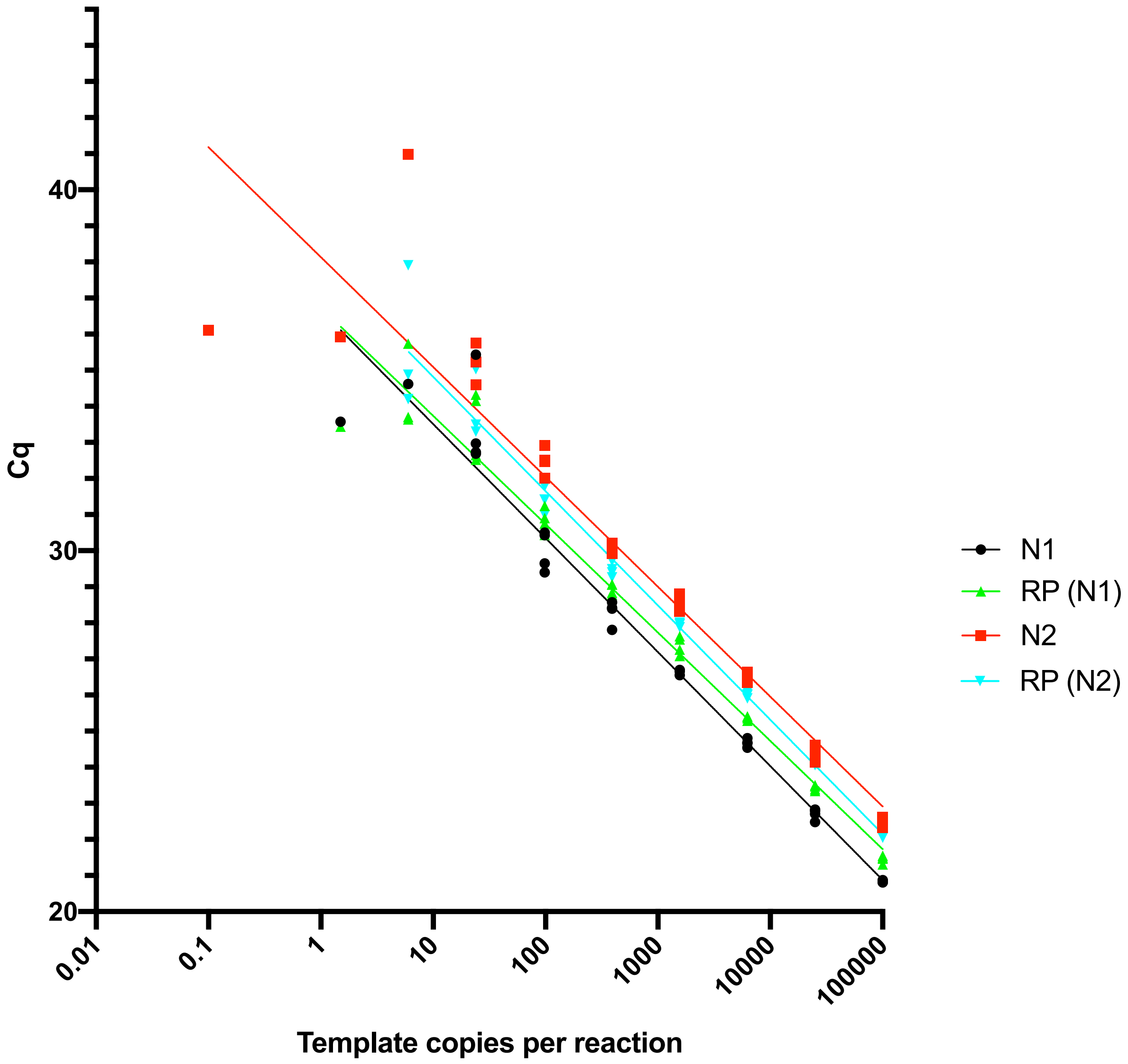


Supplementary Figure 2. **RT-qPCR sensitivity plot.** As described in Materials and Methods the limit of detection and linearity of the SARS-CoV-2 virus assay was tested using the control plasmids IDT 2019 nCoV_N_Positive Control and IDT 2019 Hs_RPP30_Positive Control. Plasmids were serially diluted from 100,000 copies/rxn down to 0.1 copies/rxn in water and RT-qPCR reactions were set up in triplicate and performed as described to determine the Cq values for each dilution.
